# Defining and phenotyping gastric abnormalities in long-term type 1 diabetes using body surface gastric mapping

**DOI:** 10.1101/2022.08.10.22278649

**Authors:** William Xu, Armen A. Gharibans, Stefan Calder, Gabriel Schamberg, Anthony Walters, Jia Jang, Chris Varghese, Daniel Carson, Charlotte Daker, Stephen Waite, Christopher N Andrews, Tim Cundy, Gregory O’Grady

## Abstract

**Objective:** To define phenotypes of gastric myoelectrical abnormalities and relation to symptoms in people with longstanding T1D, compared to matched healthy controls, using a novel non-invasive body surface gastric mapping (BSGM) device.

**Research design and methods:** BSGM was performed on people with T1D of >10 years duration and matched controls, employing Gastric Alimetry (Alimetry, New Zealand), comprising a high-resolution 64-channel array, validated symptom logging App, and wearable reader.

**Results:** 32 people with T1D were recruited (15 with a high symptom burden), and 32 controls. Those with symptoms showed more unstable gastric myoelectrical activity, (Gastric Alimetry Rhythm Index 0.39 vs 0.51, p=0.017; and lower average spatial covariance 0.48 vs 0.51, p=0.009) compared with controls. Those with T1D and symptoms also had higher prevalence of peripheral neuropathy (67% vs 6%, p=0.001), anxiety/depression diagnoses (27% vs 0%, p=0.001), and mean HbA1c levels (76 vs 56 mmol/mol, p<0.001). BSGM defined distinct phenotypes in participants including those with markedly unstable gastric rhythms (4/32, 12.5%), and abnormally high gastric frequencies (10/32, 31%). Deviation in gastric frequency was positively correlated with symptoms of bloating, upper gut pain, nausea and vomiting, and fullness and early satiation (r>0.35, p<0.05)

**Conclusion:** Gastroduodenal symptoms in people with longstanding T1D correlate with gastric myoelectrical abnormalities on BSGM evaluation, in addition to glycemic control, psychological comorbidities, and peripheral neuropathy. BSGM using the Gastric Alimetry device identified a range of myoelectrical phenotypes, representing both myogenic and neurogenic mechanisms, which represent targets for diagnosis, monitoring and therapy.

## Introduction

Gastric symptoms are common in people with type 1 diabetes (T1D), impairing quality of life, contributing to nutritional compromise, and complicating glycemic control.^1^ Symptoms such as early satiety and excessive fullness, nausea and vomiting, bloating, and distension can be severe and refractory to medical therapy.^2,3^

These symptoms are commonly attributed to diabetic gastroparesis, a disorder of gastric emptying which affects about 5% of people with T1D.^1^ However, people with diabetes can have disruptive symptoms without gastric emptying abnormalities or may have gastrointestinal symptoms of non-gastric origin, so the more inclusive terminology of ‘diabetic gastroenteropathy’ has been suggested.^2,4^

Diabetic gastroenteropathy is multifactorial. Autonomic or enteric neuropathy, immediate glycemic changes, brain-gut axis dysfunction, abnormal gastric emptying, and impaired fundic accommodation have been identified as contributing factors.^3^ Degradation of interstitial cell of Cajal (ICC) networks and disordered slow wave activity have also been implicated in disease pathophysiology, with decreased ICC and rhythm disturbances found in many patients with refractory diabetic gastroparesis and chronic nausea and vomiting syndrome. ^5–7^

Previous studies using electrogastrography (EGG) have shown frequency and rhythm abnormalities in symptomatic T1D subjects.^8,9^ However, the use of EGG is largely restricted to research and has not achieved widespread clinical adoption owing to technical limitations including high sensitivity to ‘noise’, lack of spatial resolution, and inability to clearly separate disease subgroups.^10^

A new technique called body surface gastric mapping (BSGM; high-resolution electrogastrography) has recently been introduced, offering a novel diagnostic tool that overcomes many of the limitations of EGG.^10–13^ BSGM employs a dense array of electrodes at the epigastric surface, and can identify novel biomarkers of slow wave stability and propagation patterns, providing superior symptom correlations and decreased sensitivity to ‘noise’.^11,12,14^ These methods may provide insight into the pathophysiology of gastric pathology in T1D, and offer clinical utility for the non-invasive evaluation of gastric function.

The aim of this study was to classify gastric myoelectrical abnormalities and their symptom correlations in people with T1D using a non-invasive BSGM medical device (Gastric Alimetry®, Alimetry™, New Zealand).

### Research design and methods

Ethical approval was gained from the Auckland Health Research Ethical Committee (AHREC: 1130) and the University of Calgary ethics committee (ID: REB19-1925) All patients provided informed written consent.

#### Study population

People aged ≥18 years with T1D of at least 10 years’ duration, with or without upper gastrointestinal symptoms were recruited in Auckland, New Zealand and Calgary, Canada. Exclusion criteria were: a history of gastric or duodenal surgery, active malignancy of the gastrointestinal tract, active gastrointestinal infection (including *H. pylori*), active neurogenic or additional endocrine disorders known to affect gastric motility (multiple sclerosis, scleroderma, Parkinson’s disease, hyperthyroidism), current pregnancy, cognitive impairment, cyclical vomiting syndrome, or cannabinoid hyperemesis.

Participants were assessed using the Rome IV criteria for chronic nausea and vomiting syndrome or functional dyspepsia to determine the presence of a significant chronic upper gastrointestinal symptom burden,^15^ and were stratified into those with and without significant symptoms based on meeting at least one of these criteria.

Participants with T1D were matched to a database of controls (110 subjects ≥18 years recruited during 2021) in a 1:1 ratio using the nearest neighbor method based on age, sex, and body mass index (BMI) with the *matchit* package.^16^ Control subjects were excluded if they had active gastrointestinal disease or symptoms, met Rome IV criteria, were taking medications altering gastrointestinal motility, or consumed regular cannabis. Tests with >50% of the duration marked as artifacts were excluded from the analysis, per the Gastric Alimetry Instructions for Use.

#### Study procedure

Basic information including age, sex and BMI was collected from each participant. Clinical variables included regular medications, duration of diabetes diagnosis, HbA1c within 1-year (mmol/mol), daily insulin use, insulin pump use, and presence of diabetic complications such as retinopathy, nephropathy, and peripheral or autonomic neuropathy. The assessment and definition of diabetes complications are detailed in the **supplementary methods**.

BSGM was performed using Gastric Alimetry (Alimetry, New Zealand), employing a high resolution 64-channel electrode array, a validated symptom logging App^17^, and a wearable reader device (**Figure S1**). The system has been extensively validated to detect gastric myoelectrical activity including spectral and spatial profiling.^13,18,19^ Further details of the test protocol are detailed in the **supplementary methods**.

Participants were requested to withhold prokinetic medications and antiemetics for at least 24 hours prior to the study, but other medications (insulin, other oral hypoglycemics, antidepressants, anxiolytics, gabapentinoids, proton pump inhibitors) were not withheld prior to the study. No subjects were taking GLP-1 analogues. Participants fasted for >6 hours prior to the study, avoiding caffeine and nicotine. After a 30 minute baseline recording, participants consumed a meal including an energy bar and a nutrient drink. Control participants consumed a 68g Clif bar (255 kcal; Clif Bar & Company, CA, USA) and 250mL of Vanilla Ensure (232 kcal), while participants with diabetes consumed either a 70g Optifast Chocolate bar (234 kcal, Nestle Healthcare Nutrition, NJ, USA) or 60g Horley’s Cookies and Cream bar (194 kcal; Horleys, Auckland, NZ) and 200mL of Diasip (208 kcal; Abbott Nutrition, IL, USA). The meal was followed by a 4 hour postprandial recording. Participants sat reclined in a chair with their legs elevated, were asked to limit movement, talking, sleeping, and refrain from touching the array, but were able to read, watch media, work on a mobile device, and mobilize for comfort breaks.

#### Patient reported outcomes

All participants completed the Patient Assessment of Upper Gastrointestinal Disorders-Symptom Severity Index (PAGI-SYM), Gastroparesis Cardinal Symptom Index (GCSI), and Patient Assessment of Upper Gastrointestinal Disorders-Quality of Life (PAGI-QOL) instruments as baseline gastrointestinal symptom severity and quality of life assessment.^20,21^ Health psychology assessments were completed using the State Trait Anxiety Inventory Short Form (STAI-SF) and Patient Health Questionnaire-2 (PHQ-2) questionnaires.^22,23^

During the test, participants were asked to rate the upper gastrointestinal symptoms of upper gut pain, nausea, bloating, heartburn, stomach burn, and excessive fullness every 15 minutes on visual analogue scales from 0 to 10 (0 indicating no symptoms; 10 indicating the worst imaginable extent of symptoms) using the validated Gastric Alimetry App.^17^ Patients rated early satiation immediately after the meal. These data were used to calculate the validated ‘Total Symptom Burden Score’.^17^ Discrete events of vomiting, belching, and reflux were recorded.

#### Continuous glucose monitoring

In a subset of people with T1D continuous glucose monitoring (CGM) was performed using Freestyle Libre Pro (Abbott Laboratories, USA) sensors during the study. Capillary blood glucose tests before the study meal, and at 30 minutes and 2 hours post-meal were undertaken to calibrate sensors (Caresens N, Pharmaco Diabetes, New Zealand).

#### BSGM analysis

Data from participants with T1D were compared to the matched healthy controls and individual data were compared to reference intervals developed from a recent study of 110 controls.^24^ Four spectral metrics reported by the Gastric Alimetry system were employed: the Gastric Alimetry Rhythm Index (GA-RI; quantifying the extent to which activity is concentrated within a narrow frequency band over time relative to the residual spectrum; abnormal <0.25), Principal Gastric Frequency (the sustained frequency associated with the most stable oscillations as defined by GA-RI; normative interval 2.65 - 3.35 cpm), BMI-adjusted amplitude (normative interval 20 - 70μV), and Fed:Fasted Amplitude Ratio (ff-AR, normative interval ≥1.08). Further explanations for each metric are detailed in the **supplementary methods** and elsewhere.^19^ Given the bimodal distribution of Principal Gastric Frequency abnormalities found in the cohort (see Results), Principal Gastric Frequency Deviation (the absolute difference of Principal Gastric Frequency from 3 cpm) was calculated to determine the magnitude of frequency abnormalities.

In addition, to assess direction of slow wave propagation, spatial propagation phase maps were generated in 15 minute epochs (**Fig S1E**).^13^ As reference intervals for BSGM spatial metrics are still under development, spatial patterns were classified by consensus (four blinded expert assessors) as antegrade, retrograde, or indeterminate (no definite slow wave propagation direction or no consensus) (**Fig S6**) according the methods of Gharibans et al.^13^ Only cases with sufficiently high GA-RI (>0.25) and a low percentage of indeterminate periods on consensus classification (<50%) were included in spatial metrics analysis. Retrograde activity for ≥20% of the study duration was defined as abnormal.

### Statistical analysis

Data are reported as the mean ± standard deviation (SD) or median and interquartile range (IQR) unless stated otherwise. Comparisons were made using one-way ANOVA with further pairwise comparisons via a post-hoc Benjamini-Hochberg correction. Fisher’s exact test was used for categorical variables. Continuous independent non-normal variables were compared using the Mann-Whitney U test. Associations between BSGM metrics, demographic data, CGM data and HbA1c, and symptoms were assessed using Pearson correlation with Benjamini-Hochberg’s correction to minimize false discovery rate. A statistical significance threshold p<0.05 was used. All analyses were performed in R version 4.0.3 (R Foundation for Statistical Computing, Vienna, Austria).

## Results

### Study population

BSGM studies were completed in 34 subjects with T1D, of which two were excluded due to excessive motion artifacts (>50% duration). The remaining 32 subjects were matched by age, sex, and BMI to 32 healthy controls (**Table 1**).

**Table 1:** Patient demographics and clinical features of diabetes.

| Variables |  | Controls | T1D - no symptoms | T1D - symptoms | Total | p |
| --- | --- | --- | --- | --- | --- | --- |
| Total N (%) |  | 32 (50) | 17 (27) | 15 (23) | 64 |  |
| Age | Mean ± SD | 47.8 ± 14.9 | 56.2 ± 14.6 | 47.5 ± 13.5 | 50.0 ± 14.8 | 0.123 |
| Sex | Female (%) | 20 (62.5) | 11 (64.7) | 10 (66.7) | 41 (64.1) | 0.960 |
| BMI | Mean ± SD | 25.3 ± 4.1 | 26.2 ± 3.6 | 25.1 ± 6.3 | 25.5 ± 4.5 | 0.772 |
| Gastroesophageal reflux disease | (%) | 0 (0) | 1 (6) | 0 (0) | 1 (2) | 0.246 |
| Hypothyroidism | (%) | 0 (0) | 4 (24) | 2 (13) | 6 (9) | **0.022** |
| Ischemic Heart Disease | (%) | 0 (0) | 2 (12) | 0 (0) | 2 (3) | 0.058 |
| Hypertension | (%) | 0 (0) | 3 (18) | 0 (0) | 3 (45) | **0.013** |
| Anxiety/Depression diagnosis | (%) | 0 (0) | 0 (0.0) | 4 (27) | 4 (6) | **0.001** |
| PTSD | (%) | 1 (3) | 1 (6) | 3 (20) | 5 (8) | 0.125 |
| Previous non-gastric abdominal surgery | (%) | 12 (39) | 8 (47) | 7 (50) | 27 (44) | 0.734 |
| Duration of diabetes diagnosis (years) | Mean ± SD | | 33 ± 15 | 30 ± 15 | 32 ± 15 | 0.576 |
| HbA1c (mmol/mol) | Mean ± SD | | 56 ± 9 | 76 ± 24 | 65 ± 20 | **0.005** |
| Insulin dose (units/day) | Mean ± SD | | 42 ± 19 | 55 ± 38 | 47 ± 29 | 0.257 |
| Insulin pump use | (%) |  | 3 (18) | 4 (27) | 7 (22) | 0.851 |
| Retinopathy - presence | (%) |  | 9 (53) | 10 (67) | 19 (59) | 0.668 |
| Retinopathy - grade | None (%) |  | 7 (41) | 6 (40) | 13 (41) | 0.986 |
|  | Mild/moder ate (%) |  | 7 (41) | 6 (40) | 13 (41) |  |
|  | Severe (%) |  | 3 (18) | 3 (20) | 6 (19) |  |
| Chronic kidney disease - grade | No CKD (%) |  | 17 (100.0) | 10 (66.7) | 27 (84.4) | 0.152 |
|  | Stage 2-3 (%) |  |  | 4 (27) | 4 (27) |  |
|  | Stage 4-5 (%) |  |  | 1 (7) | 1 (3) |  |
| Peripheral neuropathy - presence | (%) |  | 1 (6) | 10 (67) | 11 (34) | **0.001** |
| Postural hypotension - presence | (%) |  | 1 (6) | 6 (40) | 7 (22) | 0.057 |

The mean age was 50 years (SD 14.8) and the majority were women (41/64, 64%). The median BMI was 25.1 kg/m^2^ (range 16.4 to 38.9) and the median duration of diabetes was 32 years (range 11 to 65). All the participants with T1D were taking insulin, apart from one symptomatic person who had an islet cell transplantation 2 years earlier. History of other medication use is detailed in **Table S1**.

### Symptoms

Of the 32 participants with T1D, 12 met the Rome IV Criteria for both chronic nausea and vomiting syndrome and functional dyspepsia, and 3 for functional dyspepsia only, indicative of significant chronic gastroduodenal symptoms in 15 subjects, while 17 subjects did not meet criteria (**Table 1**). Those with gastroduodenal symptoms had higher HbA1c levels compared to those without symptoms (76 ± 24 mmol/mol vs 56 ± 9 mmol/mol; p=0.005) and an increased prevalence of peripheral neuropathy (67% vs 6%; p=0.001). Those with symptoms also had higher prevalence of anxiety and/or depression clinical diagnoses (27% symptoms vs 0% no symptoms vs 0% controls; p=0.01, **Table 1**).

Participants with T1D and gastrointestinal symptoms had higher PAGI-SYM (2.35 (1.18 to 2.70) symptoms vs 0.15 (0.05 to 0.25) no symptoms vs 0.10 (0.00 to 0.31) controls; p<0.001) and GCSI scores (2.89 (1.39 to 3.44) symptoms vs 0.11 (0.00 to 0.44) no symptoms vs 0.00 (0.00 to 0.22) controls; p<0.001, **Figure 1, Table S2**). Symptomatic participants had reduced quality of life as measured by the PAGI-QoL score (2.31 (1.26 to 3.43) symptoms vs 0.20 (0.03 to 0.43) no symptoms vs 0.12 (0.00 to 0.32) controls; p<0.001, **Table S2**), and demonstrated higher STAI-SF and PHQ-2 scores (all p<0.05; **Figure 1, Table S2**).

**Figure 1:**
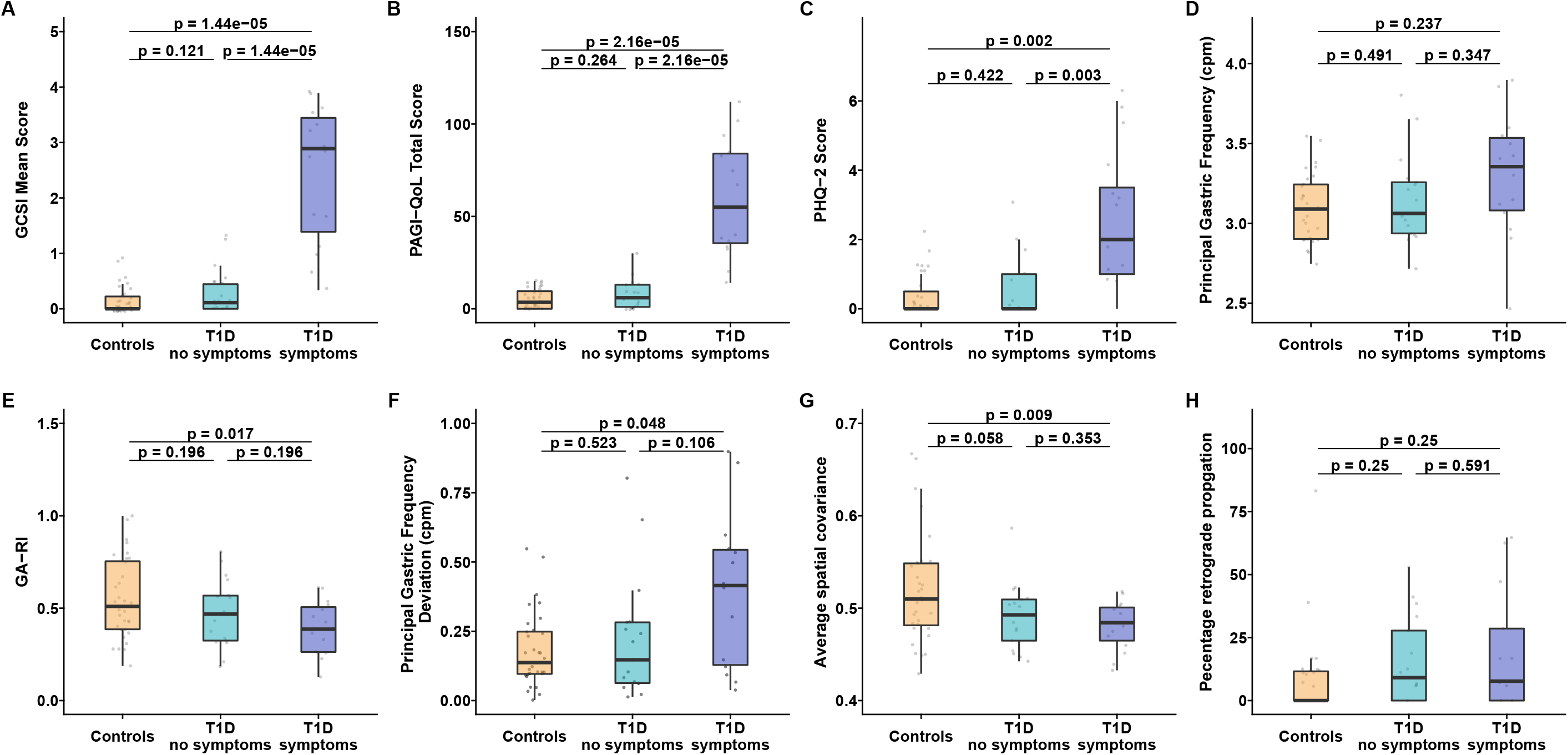
BSGM metric differences between subjects with T1D and controls. Box plots of symptoms and metrics across controls, those with type 1 diabetes mellitus without high symptom burden (T1D no symptoms), and those with T1D and a high symptom burden (T1D symptoms; as defined by meeting Rome IV Criteria for chronic nausea and/or vomiting syndrome or functional dyspepsia). GCSI, Gastroparesis Cardinal Symptom Index; PAGI-QoL, Patient Assessment of Upper Gastrointestinal Disorders–Quality of Life Index; PHQ-2, Patient Health Questionnaire-2; GA-RI, Gastric Alimetry Rhythm Index.

### BSGM: Whole Cohort Analysis

BSGM metrics between patient groups are reported in **Figure 1** and **Table S2**, and technical metrics of the test in **Table S3**. One person with T1D without symptoms required additional oral glucose at the 2 hour post-meal stage for threatened hypoglycemia and had a shortened BSGM test, while one person with symptoms had device disconnection and data loss for one hour of the test. All except for one symptomatic person finished ≥50% of the study meal, which is shown as providing a sufficient stimulus to generate reliable meal response metrics,^14^ (**Table S3**).

#### Rhythm, frequency and stability

Participants with T1D who had symptoms showed marked abnormalities in gastric slow wave stability compared with healthy controls, with reduced GA-RI (0.39 (0.26 to 0.51) vs 0.51 (0.39 to 0.75); p=0.017), higher Principal Gastric Frequency Deviation (0.41cpm (0.13 to 0.54) vs 0.14cpm (0.10 to 0.25); p=0.048), and reduced spatial organization of gastric wavefronts (average ‘spatial covariance’ 0.48 (0.46 to 0.50) vs 0.51 (0.48 to 0.55); p=0.009) (**Figure 1, Table S2**). In contrast, participants with T1D without symptoms showed minor or no difference compared to healthy controls in GA-RI (0.47 (0.32 to 0.57) vs 0.51 (0.39 to 0.75), p=0.203), Principal Gastric Frequency Deviation (0.15cpm (0.06 to 0.28) vs 0.14cpm (0.10 to 0.25) in controls; p=0.510), and spatial organization of wavefronts (0.49 (0.46 to 0.51) vs 0.51 (0.48 to 0.55); p=0.058) (**Table S2**).

#### Amplitude

There were no differences in BMI-adjusted amplitude between those with T1D with or without gastric symptoms (median 40.5 µV (25.7 to 47.8) symptoms vs 35.0 µV (33.0 to 40.9) no symptoms vs 33.3 (27.1 to 50.0) controls; p=0.975). Similarly there were no differences in the Fed:Fasted Amplitude Ratios (1.62 (1.51 to 2.13) symptoms vs 1.80 (1.36 to 2.26) no symptoms vs 1.87 (1.47 to 2.22) controls; p=0.927) (**Table S2**).

#### Relationship between BSGM metrics, symptom burden, and blood glucose levels

Overall median blood glucose level averaged over the 30 minute pre-study period was 7.8 mmol/L (6.6 to 10.1) in those with CGM monitoring (n=20) and median levels averaged over the post-meal period were 9.3 mmol/L (8.5 to 12.0) (n=21) (**Table S2**). Blood glucose levels over the post meal period stratified by symptom group are displayed in **Figure S2**.

Correlations between symptoms, blood glucose levels, and BSGM metrics after Benjamini-Hochberg’s corrections are displayed in **Figure 2G**. HbA1c, PHQ-2, and STAI-SF scores were positively correlated with GCSI and symptoms such as bloating or nausea and vomiting (r>0.5; p<0.05). Principal Gastric Frequency Deviation was positively correlated with GCSI score and symptoms of bloating, upper gut pain, nausea and vomiting, and fullness and satiety (r>0.35; p<0.05)

**Figure 2:**
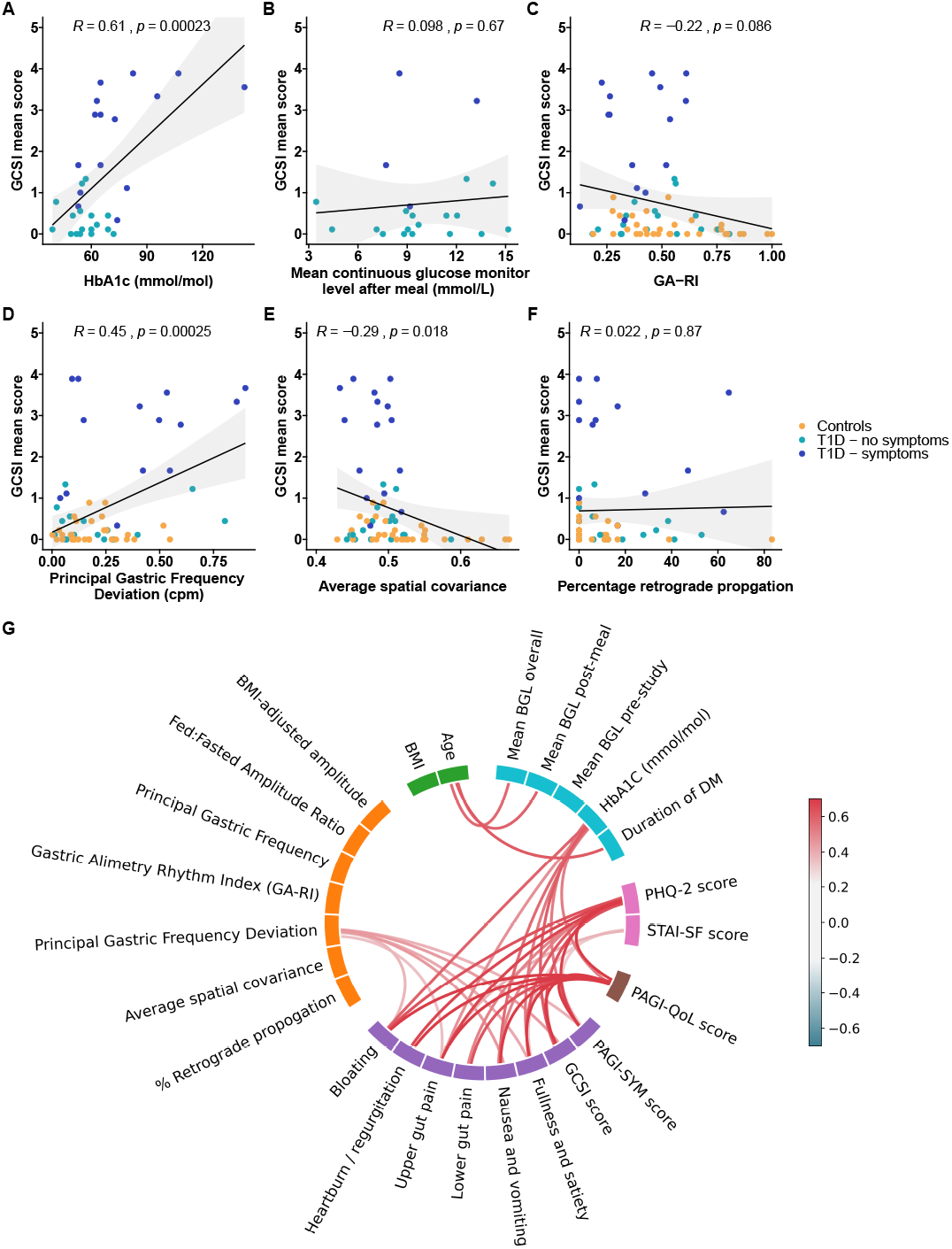
Correlations between blood glucose, BSGM metrics, and symptoms. A-F) Correlation plots of symptoms and metrics across controls, those with type 1 diabetes mellitus without high symptom burden (T1D no symptoms), and those with T1D and a high symptom burden (T1D symptoms; as defined by meeting Rome IV Criteria for chronic nausea and vomiting syndrome and/or functional dyspepsia (normal)). Total symptom burden: sum of all symptom scores measured by the validated Alimetry app averaged over the post-meal phase. Unadjusted p-values shown. CGM data was available for analysis in 20/31 diabetic participants (n=16 minimal symptoms, n=4 significant upper GI symptoms). G) Wheel plot showing correlations between demographics, BSGM metrics, symptoms as measured by the PAGI-SYM subscales, PAGI-SYM quality of life, health psychology metrics, and diabetes factors. Only statistically significant correlations between categories are shown (p<0.05 after Benjamini-Hochberg correction). BSGM, body surface gastric mapping; GCSI, gastroparesis cardinal symptom index; PAGI-SYM, Patient Assessment of Upper Gastrointestinal Disorders–Symptom Severity Index; PAGI-QoL, Patient Assessment of Upper Gastrointestinal Disorders-Quality of Life; STAI-SF, State-Trait Anxiety Inventory Short Form; PHQ-2, Patient Health Questionnaire-2; DM, diabetes mellitus; BGL, blood glucose level; GA-RI, Gastric Alimetry Rhythm Index.

Blood glucose levels were related to BMI-adjusted Amplitude (r=0.49; p=0.025), and Principal Gastric Frequency (r=0.4; p=0.084; **Fig 3H**) on univariate analysis. However, after adjustment for multiple comparisons these results were not statistically significant (**Figure 2G**).

**Figure 3:**
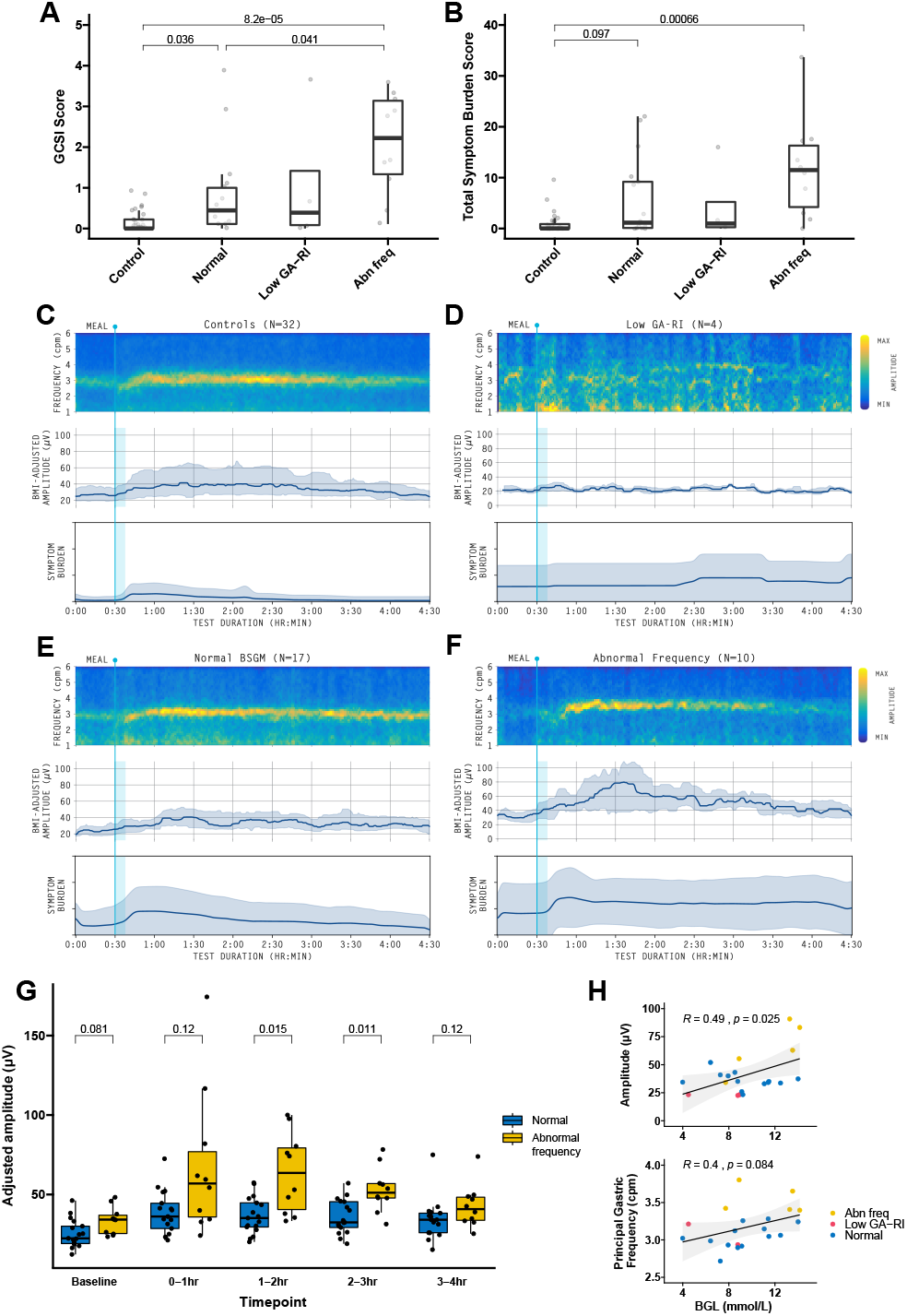
BSGM Phenotypes. Symptom severity measured by A) GCSI score and B) Total Symptom Burden Score between healthy controls and T1D participants based on BSGM classification.^17^ C) Averaged BSGM spectral plots (frequency-amplitude graphs), showing Principal Gastric Frequencies on a scale from low power (dark blue) to high power (bright yellow), indicating gastric meal responses and rhythm between different groups. The mealtime is indicated by a vertical bar at 30 min. C) and E) Normal spectral plots show a clear meal response (post-prandial power increase), a consistent and sustained frequency band, and a regular gastric rhythm. D) Low Gastric Alimetry Rhythm Index (GA-RI) cases lacked these features. F) In a large subgroup of participants, abnormal frequencies were shown, with predominantly high frequency cases and one low frequency case (abn freq). Accompanying BMI-adjusted amplitude (median and IQR) and symptom burden plots (mean and SD) over time shown. A T1D participant with isolated abnormal amplitude is presented in **Figure S5**. G) BMI-adjusted amplitude compared between abnormal frequency and normal BSGM subgroups over time. H) Scatter plot showing relationship between adjusted amplitude, Principal Gastric Frequency, and blood glucose levels.

Scatter plots for pairwise comparisons between glycemic levels, Principal Gastric Frequency Deviation, average ‘spatial covariance’, and symptom scores are displayed in **Figure 2A-F**.

### BSGM: Phenotype Analysis

A flowchart detailing the classification of spectrograms is presented in **Figure S3**. BSGM spectral data falling outside the reference intervals was more common in people with T1D and symptoms (67% vs 29% without symptoms; p=0.074).

Using reference interval data, participants with T1D were grouped into phenotypes. Out of the 32 people with T1D, four (12.5%) were classified with unstable rhythms (GA-RI <0.25), and a further 10 subjects with abnormal Principal Gastric Frequencies despite normal GA-RI (>3.35 or <2.65cpm; GA-RI ≥ 0.25; 31%).^19,24^ Nine subjects (28.1%) had high frequencies (>3.35cpm) and one symptomatic subject (3.1%) had low frequency activity (2.5cpm). Three people with T1D (9%) had abnormally high BMI-adjusted amplitudes (>70μV), with this being the only abnormal BSGM finding in one (3%). One person with T1D (3%) with an unstable GA-RI also had a low Fed:Fasted Amplitude Ratio of 0.80 (reference interval ≥1.08). A total of 17/32 subjects with T1D (31%) had no abnormality detected on spectral analysis (**Figure S3**). Averaged spectrograms, amplitude plots, and symptom burden during the study across specific phenotypes are displayed in **Figure 3A-3F**.

On spatial pattern analysis, data from eight individuals were excluded as >50% of their test duration was classified as spatially indeterminate (3 symptomatic subjects, 2 subjects without symptoms, 3 controls). There was no overall group-level statistical difference for time spent in retrograde propagation between subjects with and without symptoms or controls (7.4% (IQR 0.0 to 19.6) vs 6.7% (0.0 to 30.6) vs 0.0% (0.0 to 12.1) respectively, p=0.318; **Figure 1, Table S2**). Three out of 12 (25%) subjects with T1D and symptoms exceeded the threshold of ≥20% retrograde activity, compared to 5/15 (33.3%) without symptoms and 2/29 (6.9%) controls (p=0.051).

#### Meal Response

People with T1D with Principal Gastric Frequency Abnormalities had higher postprandial BMI-adjusted amplitudes than those with normal BSGM (1-2hr: 63.6 µV (40.5 to 79.3) high frequency vs 35.2 (29.5 to 44.8) normal BSGM; p=0.015; 2-3hr: 51.1 µV (47.8 to 56.9) vs 32.5 (29.2 to 45.5); p=0.011; **Figure 3G**). Amplitude was more closely correlated with BGL in the abnormal frequency and normal BSGM subgroups compared to the low GA-RI subgroup (**Figure S4**).

### Sensitivity analysis

Sensitivity analysis was conducted excluding one subject with symptoms who did not withhold domperidone on the day of the study. No differences to the main BSGM or spectral and spatial analysis were demonstrated.

### Safety

No participants experienced a serious adverse event. Mild transient skin redness or rash (n=10) and itch (n=5) lasting <48 hours occurred in a subset of participants with spontaneous resolution. On a scale of 0-10 (0 indicating unlikely, 10 highly likely), participants were likely to recommend the test (median 10; IQR (8 to 10)).

## Conclusions

This study applied a new non-invasive medical device (BSGM; Gastric Alimetry) to characterize gastric myoelectrical abnormalities in people with longstanding T1D. Disrupted gastric activity represented by unstable rhythms, abnormal frequencies, and abnormal spatial propagation was shown to occur more regularly in symptomatic patients, with significant correlations to symptoms. Glycemic control, psychological comorbidities, and peripheral neuropathy were also correlated with symptoms, reinforcing the multifactorial nature of diabetic gastroenteropathy. Most significantly, the use of Gastric Alimetry enabled phenotyping of T1D gastropathy at the individual patient level based on novel spectral metrics and their reference intervals, representing distinct targets for diagnosis, monitoring and therapy. Key phenotypes included a group with high Principal Gastric Frequency without gastric rhythm instability (28% of participants) associated with high symptom burden, distinct from a smaller group of participants with highly unstable gastric rhythms (i.e. low GA-RI) and lower symptom burdens.

Gastric dysrhythmias are postulated as a key mechanism in diabetic gastropathy, and have been consistently demonstrated in people with T1D suffering gastric symptoms.^8,12,25^ However, gastric dysrhythmias are heterogeneous.^26^ Invasive high-resolution serosal mapping studies have defined specific physiological categories of dysrhythmia, including stable and unstable ectopic pacemaker activity at normal and abnormal frequencies, and aberrant spatial conduction pathways, which explain the abnormalities seen non-invasively in the present study.^6,7^ Legacy EGG studies showed that people with T1D experienced lower time durations in normal gastric rhythms.^8,27^ However, such results required cautious interpretation as abnormal frequencies measured by legacy EGG could include biologically implausible gastric frequencies (e.g. <1.5 or >5cpm), and may conflate artifacts with stable gastric activity,^28^ such that they could not provide independent measures of frequency and rhythm.^19^ BSGM provides substantially improved capture of gastric myoelectrical activity, and has metrics that overcome these limitations including distinct measures of frequency and rhythm,^19^ allowing a more accurate and specific non-invasive assessment of gastric myoelectrical abnormalities in T1D.

The vagus nerve plays an important role in regulating ICC frequency,^28^ and stable high gastric frequencies have been variably demonstrated after vagotomy.^29,30^ The persistent high frequency phenotype observed in our symptomatic group is plausibly a manifestation of vagal dysfunction. Vagal nerve pathology in diabetic gastropathy was first postulated in the earliest observations of diabetic gastroparesis by Kassander,^31^ and vagal nerve abnormalities such as reduced myelinated fiber density have been demonstrated in humans, albeit inconsistently.^32–34^ Low vagal nerve tone has also been associated with impaired gastric accommodation in T1D.^35^ The strong association of peripheral neuropathy and gastric symptoms, shown both in this study and others, reinforces the presence of a neurogenic mechanism in diabetic gastropathy.^36^ Vagal neuropathy may be one unifying pathophysiological mechanism causing frequency disturbance, impaired gastric sensori-motor dysfunction, reduced accommodation, and symptoms observed in T1D.^2^ Notably, autonomic neuropathy in diabetes often co-occurs with enteric nervous system, ICC and smooth muscle pathology, such that additional factors could also be contributory in any individual patient.^37^

A smaller subgroup in this study showed a BSGM phenotype with weak, irregular, and unstable gastric myoelectrical activity (low GA-RI) accompanied by impaired meal responses. Such abnormalities are considered to represent gastric neuromuscular abnormalities,^14^ and may be consistent with underlying depletion and damage to ICC networks in diabetic gastropathy,^5,6^ which are known to contribute to abnormal slow wave dynamics.^6,7^ Unstable myoelectrical disturbances have recently been described in patients with nausea and vomiting syndromes.^14^ Grover and colleagues previously identified ICC reductions in 50% of patients with confirmed diabetic gastroparesis on histological analysis, and correlations with gastric emptying data would be of interest in future.^5^ In our study, a number of people with T1D also displayed high amounts of retrograde gastric slow wave activity, which was less frequently observed in healthy controls. Sustained retrograde activity has recently been revealed to be an abnormal feature in gastric disorders and symptom correlations have been demonstrated,^6,7,12,25^ but further work is needed to understand this novel finding and its clinical significance in T1D.

Hyperglycemia is known to influence gastric electrophysiology and motility, potentially inducing gastric dysrhythmia and antral hypomotility. Glucose measurements performed in the current study showed an association between BGL with Principal Gastric Frequency on univariate analysis. However, this did not persist after correction for multiple comparisons. Legacy EGG studies have attributed the presence of tachygastria to acute hyperglycemia,^27,38^ but also have found that tachygastria persists during symptoms despite euglycemia maintenance.^27^ More recent invasive mucosal and high-resolution serosal mapping techniques have revealed that hyperglycemia-induced dysrhythmias are typically transient, occurring during acute changes in blood glucose, and are characterized by spatial and rhythmic disorganization rather than sustained stable high frequency activity seen in this study.^39,40^ Further studies could evaluate blood glucose and its association with gastric abnormalities over longer time periods using emerging wearable devices.^41^

Subjects in the abnormal frequency cohort also had higher gastric amplitudes than those with normal BSGM, and an association between adjusted amplitude and blood glucose levels was observed. One hypothesis is that vagal nerve dysfunction induces rapid gastric emptying,^42^ which worsens glycemic control and raises BGL.^1^ However, further studies on this novel finding are needed to determine the direction of causality.

We found that several participants with diabetes had normal BSGM testing, with a minority showing normal gastric activity despite the presence of symptoms. Their symptoms might be mediated through other mechanisms such as visceral hypersensitivity or disorders of gut-brain interaction.^2^ Additionally, one person had abnormally high gastric amplitude of unknown significance.

The association between psychological comorbidity and symptom burden is not surprising. Associations between the complications of diabetes, poor glycemic control, and psychological comorbidities are well known,^43^ including the association between poor glycemic control and gastric symptoms.^44^ Our findings, like others, underscore that psychological comorbidity is common, and may contribute to gastrointestinal sensorimotor dysfunction in diabetes independently through the gut-brain axis, indicating another area of potential therapeutic importance.^2^

People with gastroduodenal symptoms in diabetes have been previously assessed with gastric transit studies, and classed as gastroparesis, rapid gastric emptying, or having normal gastric emptying based on transit testing.^45^ Gastric emptying remains an important assessment modality in diabetic gastropathy, but has come under recent controversy due to the inconsistent relationship between emptying rate and symptoms, the lack of resolution of symptoms despite gastric emptying improvements, and the variable repeatability of gastric emptying results over time.^4,46,47^ While this study focused on gastric myoelectrical activity and emptying testing was not performed, future studies using both BSGM and gastric emptying simultaneously would be of interest to evaluate how BSGM phenotypes and gastric emptying compare.

Several limitations of this study are acknowledged. Although most participants had blood glucose level monitoring to account for the effect that hyperglycemia has on stomach motility, controls and some participants with T1D did not receive CGMs due to device availability. One subject with T1D did not have their prokinetic medication withheld, however, a sensitivity analysis reviewed no significant differences. Although there were differences in the meals given to participants with diabetes, the caloric load and volume of the meals remained comparable. Furthermore, while spectrograms were interpreted using validated reference intervals, reference values for emerging spatial metrics are still being developed, meaning that a consensus-based classification was required.

In conclusion, gastroduodenal symptoms in people with T1D correlate with myoelectrical abnormalities detected with a novel BSGM medical device. Other factors associated with symptoms included glycemic control, psychological comorbidities, and peripheral neuropathy. Most notably, BSGM using the Gastric Alimetry was able to identify separate disease phenotypes in T1D characterized by abnormal rhythm stability and abnormal gastric frequencies, representing distinct targets for diagnosis, monitoring and therapy.

## Data Availability

All data produced in the present study are available upon reasonable request to the authors

## Acknowledgements

We thank Gen Johnson, India Wallace, Yasmin Alsagoff, Sonia Binden, Lynn Wilsack, and Renata Rehak for their outstanding assistance with participant recruitment, and to all our study participants for enabling this work.

