## supplementary methods for "Defining and phenotyping gastric abnormalities in long-term type 1 diabetes using body surface gastric mapping"

*Phenotyping gastric abnormalities in type 1 diabetes*

**Authors:** William Xu[1], Armen A. Gharibans[1,2,3], Stefan Calder[1,2], Gabriel Schamberg[1,2], Anthony Walters[4], Jia Jang[1], Chris Varghese[1], Daniel Carson[1], Charlotte Daker[2], Stephen Waite[2], Christopher N Andrews[2,5], Tim Cundy[6], Gregory O'Grady[1,2,3]

##### Affiliations

1. Department of Surgery, the University of Auckland, New Zealand
2. Alimetry Ltd, Auckland, New Zealand
3. Auckland Bioengineering Institute, The University of Auckland, New Zealand
4. Liggins Institute, University of Auckland, New Zealand
5. Dept of Gastroenterology, University of Calgary, Canada
6. Department of Medicine, University of Auckland, New Zealand

### Supplementary materials

### Supplementary methods

#### Assessment of diabetes complications

*Retinopathy*

Retinopathy was diagnosed based on ophthalmology clinical reports and classified as either no disease, mild to moderate diabetic retinopathy, or severe disease (severe non-proliferative or pre-proliferative diabetic retinopathy or proliferative retinopathy).

*Nephropathy and neuropathy*

Nephropathy was graded based on estimated glomerular filtration rate (eGFR) thresholds and clinical records (1). A diagnosis of peripheral neuropathy was made based on clinical electronic records as documented by endocrinology or neurology consult notes.

*Cardiovascular and autonomic dysfunction*

Patients had a diagnosis of hypertension and ischemic heart disease recorded from clinical records. Orthostatic hypotension was assessed as a screening tool for sympathetic nervous system dysfunction and was defined as >20mmHg drop in systolic blood pressure or >10mmg drop in diastolic after 5 minutes supine and 1 minute of upright standing (2).

#### Gastric Alimetry® System

*Section adapted from Gharibans et al. (3)*

Gastric Alimetry® is a novel medical device custom designed for BSGM. The device consists of an HR Array, wearable Reader, Dock, iPadOS App for setup and symptom logging, and cloud-based analytics and reporting platform **(Fig. S1A**). Key design considerations for each component are detailed below.

- Array. The Gastric Alimetry Array™ includes 66 pre-gelled Ag/AgCl electrodes (8×8 grid +2 reference; inter-electrode spacing 20 mm), covering an area of 21×16 cm (196 cm2) **(Fig. S1B**). This Array™ specification was designed to overlie the majority of the stomach's area in >95% of cases, which is important because gastric position is highly variable, and the weak gastric signal strength diminishes exponentially from source (4–6). Each Array is single-use, being screen printed using conductive inks on a single, flexible, thermoplastic polyurethane (TPU), with an overlying peel-and-stick adhesive layer that enables rapid setup and removal (7).
- Reader. The Alimetry Reader™ incorporates custom-designed electronics specifically tuned for gastric electrophysiology (**Fig. S1B**). Signals are acquired at 250 Hz, then amplified and digitized by low-noise programmable gain amplifiers, with each input compared against a common reference electrode to provide unipolar recordings, while movement artifacts are registered by an onboard accelerometer. The Reader attaches to the Array using a custom board-to-board connector design that eliminates all cabling to enable unimpeded wearability and facilitate ease of cleaning.
- Dock. The Alimetry Dock™ is used for charging and storage of the Reader, and accurate alignment of the Array during setup (**Fig. S1B**).

- App. The Gastric Alimetry App™ runs on an iPad mini (Apple, Cupertino, CA), and is used for device setup, data transfers, and to capture patient-reported symptom data during testing. Guided setup in the App includes an Array positioning step that tailors placement per individual patient biometrics, to further enhance accurate positioning over the stomach (7). Patients log symptoms every 15 minutes via a digital interface employing pictograms (**Fig. S1A**), which has been validated to enable reliable capture of patient symptom data in association with a standard meal with excellent compliance (8) This system therefore enables precise temporal correlations between patient symptom profiles with electrophysiological data.
- Cloud / Portal. Test data is transmitted to a HIPAA-compliant cloud server at the conclusion of each test. A proprietary algorithm automatically filters and analyzes raw myoelectrical signals to generate a report, including key metrics and data visualizations (detailed below), which are accessible via a secure online portal. Filtering methods are based on a previously validated scheme by Gharibans et al, accounting for accelerometer data (9).

#### Spatial and spectral data analytics

##### *SPECTRAL ANALYSIS*

BSGM spectrograms visualize the bioelectrical slow waves that coordinate gastric activity, as well as amplitude changes which represent meal-responses (**Fig. S1**). Four revised BSGM metrics have recently been developed to overcome several pitfalls of traditional EGG metrics (10), providing for accuracy improvements on top of the other advantages of BSGM over traditional EGG, including greater coverage over the stomach area to account anatomical variation of stomach location, a larger number of electrodes, modern bio-amplifiers, and validated signal processing techniques to decrease noise including signals from competing biological sources (3,7).

The revised BSGM metrics include BMI-Adjusted Amplitude, Principal Gastric Frequency, Gastric-Alimetry Stability Index (GA-RI), and Fed:Fasted Amplitude Ratio. Detailed descriptions of metrics are presented in **Supplementary Table 4**. Normative ranges for these revised BSGM spectral metrics were developed from a cohort of 110 health controls (11).

Patient phenotyping, as described in the methods, was subsequently completed by comparing individual subject-level data with these reference ranges.

##### *SPATIAL ANALYSIS*

The high-resolution electrode array was used to derive metrics to detect abnormal gastric slow wave activation patterns (12–15). The spatial metrics assessed in this study included 'average spatial covariance' and the percentage duration of retrograde wave propagation during the Gastric Alimetry test (3,7,14).

Average spatial covariance was defined by the average absolute value of the covariance between pairs of adjacent electrodes computed over the course of a Gastric Alimetry test.

Direction of slow wave propagation was determined by visually inspecting slow wave propagation animations averaged over 15 minute epochs per the methods of Gharibans et al (7). Example visualizations of phase map animations are displayed in **Figure S6.**

#### Strobe checklist

This study was reported according to the STROBE statement (16).

### Supplementary Tables and Figures

#### Table S1: Medication and nicotine use

One participant with T1D did not withhold their domperidone on the study day. One participant with T1D was on pancreatic enzyme replacement taking Creon.

* Denotes at least once in the last 3 months but not in the last 48 hours prior to the study

| **Variable** | | **Controls** | **T1D - no symptoms** | **T1D - symptoms** | **Total** | **p** |
|---|---|---|---|---|---|---|
| Total N (%) | | 32 (50) | 17 (27) | 15 (23) | 64 | |
| Nicotine Use* | (%) | 0 (0) | 0 (0) | 2 (13) | 2 (3) | **0.034** |
| Cannabis use* | (%) | 2 (6) | 0 (0) | 1 (7) | 3 (5) | 0.565 |
| SLGT2 Inhibitor use | (%) | 0 (0) | 2 (12) | 2 (13) | 4 (6) | 0.116 |
| Metformin use | (%) | 0 (0) | 0 (0) | 1 (7) | 1 (2) | 0.190 |
| Prokinetic use | (%) | 0 (0) | 0 (0) | 5 (33) | 5 (8) | **<0.001** |
| Pain neuromodulator use | (%) | 0 (0) | 1 (6) | 5 (33) | 6 (9) | **0.001** |
| Opioid use | (%) | 0 (0) | 0 (0) | 2 (13) | 2 (3) | **0.034** |
| Selective serotonin reuptake inhibitor, benzodiazepine use | (%) | 0 (0) | 1 (6) | 4 (27) | 5 (8) | **0.006** |
| PPI use | (%) | 3 (9) | 4 (24) | 6 (40) | 13 (20) | **0.048** |
| Antiemetic use | (%) | 0 (0) | 0 (0) | 2 (13) | 2 (3) | **0.034** |
| Laxative use | (%) | 0 (0) | 0 (0) | 1 (7) | 1 (2) | 0.190 |

#### Table S2: BSGM metrics, symptom, and quality of life data between controls and T1D patients with and without symptoms

P-values with Benjamini-Hochberg's corrections for multiple comparisons displayed. Sx, symptoms

| **Variable** | | **Controls** | **T1D - no symptoms** | **T1D - symptoms** | **Total** | **p-value** | | |
|---|---|---|---|---|---|---|---|---|
| | | | | | | **T1D - no sx vs Controls** | **T1D – sx vs Controls** | **T1D - sx -T1D vs no sx** |
| Total N (%) | | 32 (50) | 17 (27) | 15 (23) | 64 | - | - | - |
| BMI-adjusted amplitude (µV) | Median (IQR) | 33.3 (27.1 to 50.0) | 35.0 (33.0 to 40.9) | 40.5 (25.7 to 47.8) | 34.9 (27.1 to 50.0) | 0.943 | 0.818 | 0.818 |
| Fed:Fasted Amplitude Ratio | Median (IQR) | 1.87 (1.47 to 2.22) | 1.80 (1.36 to 2.26) | 1.62 (1.51 to 2.13) | 1.82 (1.42 to 2.25) | 0.780 | 0.780 | 0.780 |
| Principal Gastric Frequency (cpm) | Median (IQR) | 3.09 (2.90 to 3.24) | 3.06 (2.94 to 3.26) | 3.35 (3.08 to 3.54) | 3.12 (2.93 to 3.30) | 0.491 | 0.237 | 0.347 |
| Gastric Alimetry - Rhythm Index (GA-RI) | Median (IQR) | 0.51 (0.39 to 0.75) | 0.47 (0.32 to 0.57) | 0.39 (0.26 to 0.51) | 0.47 (0.34 to 0.61) | 0.196 | **0.017** | 0.196 |
| Principal Gastric Frequency Deviation | Median (IQR) | 0.14 (0.10 to 0.25) | 0.15 (0.06 to 0.28) | 0.41 (0.13 to 0.54) | 0.17 (0.09 to 0.32) | 0.523 | **0.048** | 0.106 |
| Average spatial covariance | Median (IQR) | 0.51 (0.48 to 0.55) | 0.49 (0.46 to 0.51) | 0.48 (0.46 to 0.50) | 0.50 (0.47 to 0.52) | 0.058 | **0.009** | 0.353 |
| Percentage time with retrograde wave propagation (%) | Median (IQR) | 0.0 (0.0 to 12.1) | 6.7 (0.0 to 30.6) | 7.4 (0.0 to 19.6) | 6.46 (0.00 to 16.67) | 0.483 | 0.483 | 0.996 |
| Mean fasting glucose (mmol/L) | Median (IQR) | NA | 7.43 (6.10 to 10.07) *n = 17* | 7.87 (7.82 to 10.65) *n = 3* | 7.82 (6.55 to 10.07) *n = 20* | - | - | 0.368 |
| Mean post-meal glucose (mmol/L) | Median (IQR) | NA | 9.33 (8.81 to 12.05) *n = 17* | 8.85 (8.32 to 10.19) *n = 4* | 9.31 (8.53 to 12.05) *n = 21* | - | - | 0.654 |
| Total Symptom Burden | Median (IQR) | 0.01 (0.00 to 1.02) | 0.34 (0.00 to 1.43) | 11.73 (8.64 to 16.68) | 0.44 (0.00 to 3.95) | 0.346 | **0.000** | **0.000** |
| GCSI | Median (IQR) | 0.00 (0.00 to 0.22) | 0.11 (0.00 to 0.44) | 2.89 (1.39 to 3.44) | 0.22 (0.00 to 0.89) | 0.121 | **0.000** | **0.000** |
| PAGI-SYM Score | Median (IQR) | 0.10 (0.00 to 0.31) | 0.15 (0.05 to 0.25) | 2.35 (1.18 to 2.70) | 0.22 (0.05 to 0.60) | 0.556 | **0.000** | **0.000** |
| PAGI-QoL Score | Median (IQR) | 0.12 (0.00 to 0.32) | 0.20 (0.03 to 0.43) | 2.31 (1.26 to 3.43) | 0.29 (0.06 to 0.53) | 0.260 | **0.000** | **0.000** |

| **Variable** | | **Controls** | **T1D - no symptoms** | **T1D - symptoms** | **Total** | **p-value** | | |
|---|---|---|---|---|---|---|---|---|
| | | | | | | **T1D - no sx vs Controls** | **T1D – sx vs Controls** | **T1D - sx -T1D vs no sx** |
| STAI-SF Score | Median (IQR) | 13.0 (10.0 to 15.5) | 24.0 (22.0 to 29.0) | 20.0 (16.5 to 31.0) | 19.0 (12.5 to 24.0) | **0.000** | **0.007** | 0.955 |
| PHQ-2 Score | Median (IQR) | 0.0 (0.0 to 0.5) | 0.0 (0.0 to 1.0) | 2.0 (1.0 to 3.5) | 0.00 (0.00 to 1.00) | 0.422 | **0.002** | **0.003** |

#### Table S3: Test quality

| Variable | | Controls | T1D - no symptoms | T1D - symptoms | Total | p-value | | |
|---|---|---|---|---|---|---|---|---|
| | | | | | | T1D - no symptoms vs Controls | T1D – symptoms vs Controls | T1D - symptoms- T1D vs no symptoms |
| Total N (%) | | 32 (50) | 17 (27) | 15 (23) | 64 | - | - | - |
| Impedance (kΩ) | Mean ± SD | 107.9 ± 78.1 | 136.7 ± 66.7 | 186.6 ± 100.1 | 134.0 ± 85.8) | 0.183 | 0.040 | 0.173 |
| Marked artifact (% duration of study) | Mean ± SD | 17.16 (11.75) | 27.81 (19.41) | 29.80 (16.68) | 22.95 (16.13) | 0.000 | 0.000 | 0.506 |
| >50% meal completion | n (%) | 32 (100) | 17 (100) | 14 (93) | 63 (98) | 0.234 | | |

**Table S4: BSGM metrics**

*Adapted from Schamberg et al. 2022 [10]*

| Metric | Description and rationale | Lower | Upper |
|---|---|---|---|
| BMI-Adjusted Amplitude (μV) | The amplitude/power (μV/dB) associated with dominant frequency in the overall spectrum is confounded by BMI. Gastric Alimetry therefore employs a conservative BMI-adjusted amplitude using a multiplicative regression. | 20 | 70 |
| Principal Gastric Frequency (cpm) | Dominant frequency calculations based on the highest average power across spectra are susceptible to transient bursts of low-frequency signal <2cpm, conflating non-gastric signals with gastric activity.[10] The 'principal gastric frequency' metric instead identifies only the frequency associated with the most stable oscillations, as measured by a distinct new stability metric (GA-RI; see below). The principal gastric frequency therefore detects the intrinsic gastric frequency, as opposed to simply calculating the frequency with the highest average power including all spectral contents whether gastric in origin or otherwise | 2.65 | 3.35 |
| Gastric Alimetry Rhythm Index (GA-RI) | Instability coefficient metrics vary in magnitude based on the dominant frequency (explained in further detail in Schamberg et al. 2022 [10]). The 'Gastric Alimetry Rhythm Index' (GA-RI), provides a measure of rhythmic gastric activity stability, by quantifying the extent to which activity is concentrated within a narrow frequency band over time relative to the residual spectrum. This improves on previous stability metrics in that it has no inherent dependence on the dominant frequency. As a result, the GA-RI enables independent assessment of the frequency and stability of gastric activity. Furthermore, the GA-RI includes a conservative BMI adjustment to account for the effect that signal attenuation has on the perceived relative strength of the gastric activity. | 0.25 | - |
| Fed:Fasted Amplitude Ratio | Amplitude increase following a meal stimulus is a characteristic of healthy gastric function. However, timing of the meal response varies. The Fed:Fasted Amplitude Ratio, instead quantifying the observed meal response by taking a ratio of the overall postprandial amplitude averaged over 4 hours to the preprandial amplitude, takes the ratio between the maximum amplitude in *any single 1-hour* across a 4-hour postprandial period to the amplitude in the preprandial period. Given that the goal of amplitude/power ratio metrics is to identify an increase in amplitude/power following meal consumption, it is important to have a metric that can quantify this increase across a cohort of subjects with significant natural variation in the timing of the meal response. | 1.08 | - |

#### Figure S1: The Gastric Alimetry system

*Adapted from Gharibans et al.* [3]

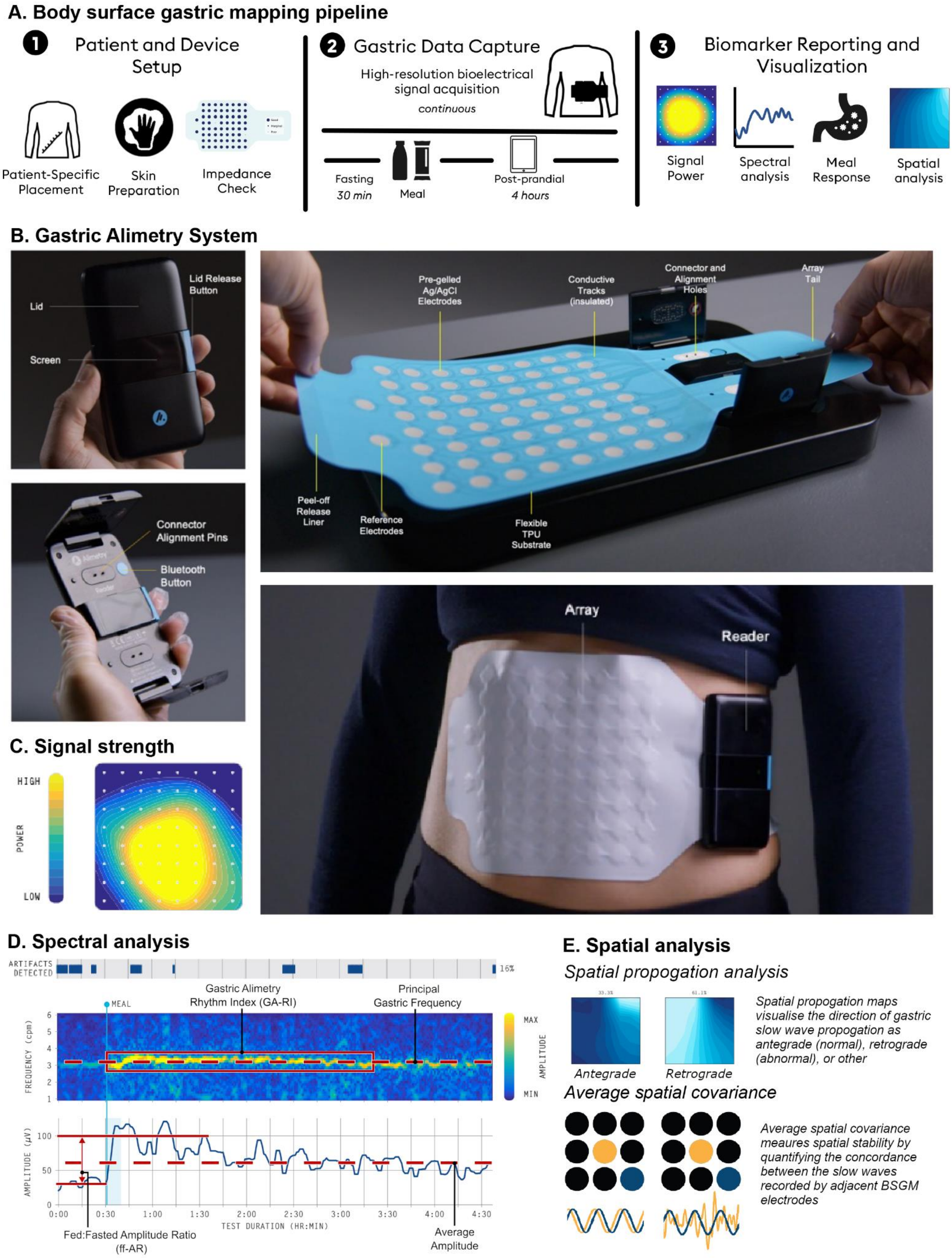

**Figure S2:**

Blood glucose levels (BGL) as measured by continuous glucose monitors during the study.

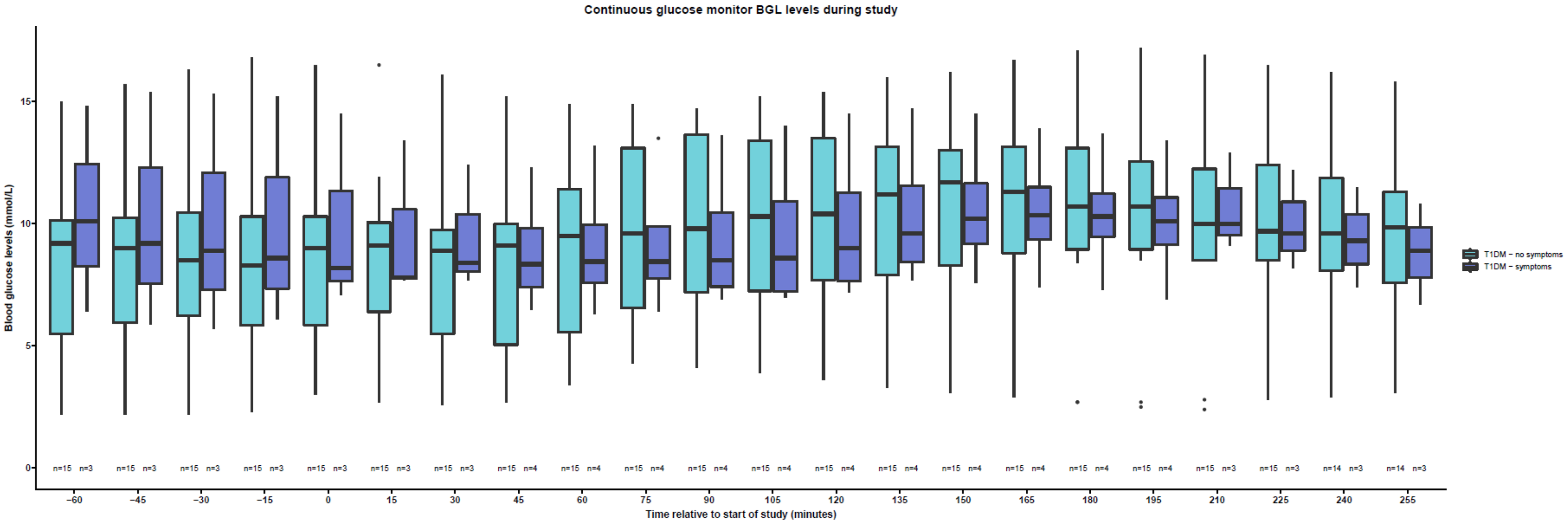

**Figure S3:**

Participant phenotype classification based on symptoms, spectrogram analysis and spatial propagation maps in this study. T1D, type 1 diabetes; sx, symptoms; CNVS, chronic nausea and vomiting syndrome; FD, functional dyspepsia; GA-RI, Gastric Alimetry Rhythm Index; PGF, principal gastric frequency; ffAR, Fed:Fasted Amplitude Ratio; SW, slow wave; BSGM, body surface gastric mapping.

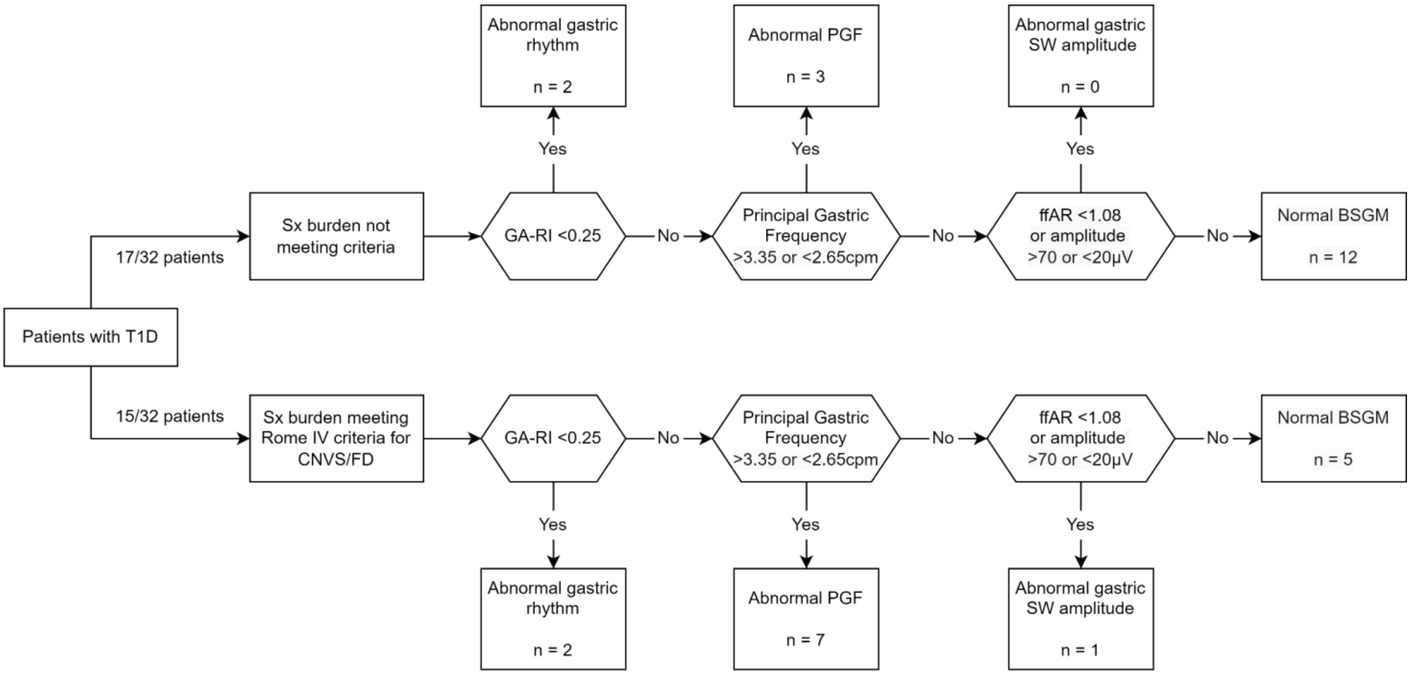

**Figure S4: Association between Principal Gastric Frequency, amplitude and blood glucose levels**

A) Within-individual pairwise Pearson R correlation coefficients for blood glucose versus amplitude averaged across phenotypes. Averaged across cohorts and compared across groups. Example plots of B) good within individual correlation between amplitude and BGL (r=0.736, p<0.001), C) poor correlation with a delayed BGL peak (r = 0.025, p=0.679), D) poor correlation with a high baseline BGL. BGL, blood glucose levels (r = -0.07, p =0.228).

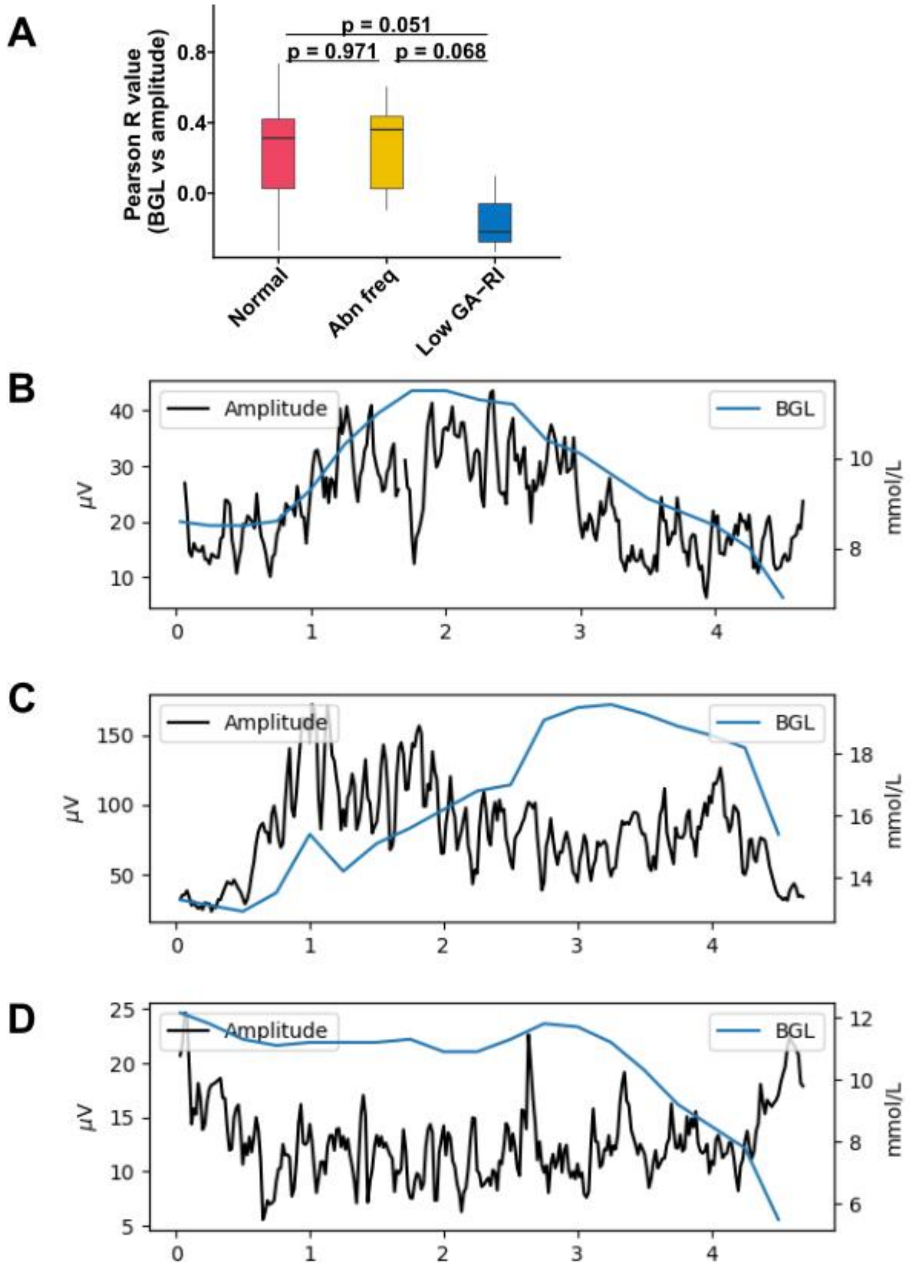

**Figure S5: BSGM of T1D patient with abnormally high amplitude**

Dark blue blocks within the spectrogram denote areas of high artifact. Note the adjusted amplitude scale relative to Figure 3.

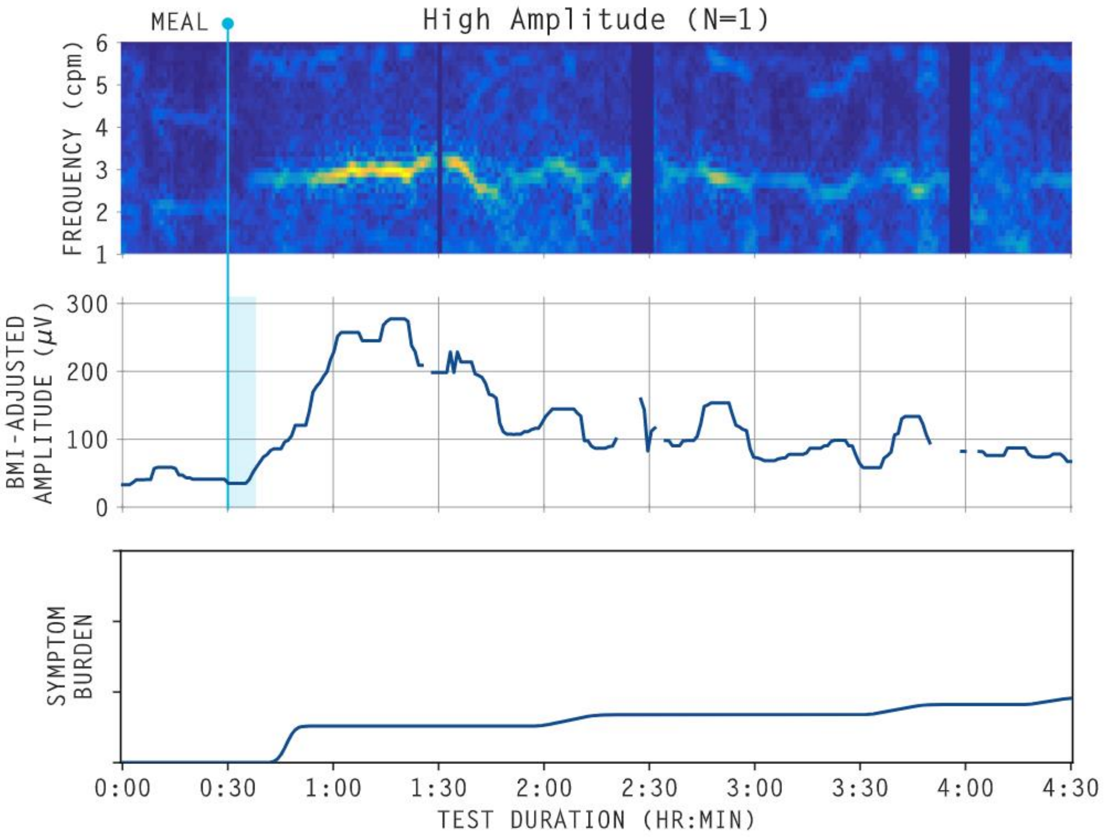

**Figure S6: Antegrade and Retrograde Slow Wave Propagation**

Spatial phase maps displaying the propagation of gastric slow waves averaged over 15 minute epochs. Frames 1 to 4 represent denote passage through time. Normal propagation is in the antegrade direction from the gastric fundus towards the gastric antrum and appears as right to left on the body surface (A).[12] Retrograde propagation in the opposite direction (B) is associated with pathological states and gastric symptoms.[14] When no clear antegrade or retrograde pattern was discernible from animations, the corresponding 15 minute epoch was marked as indeterminate.

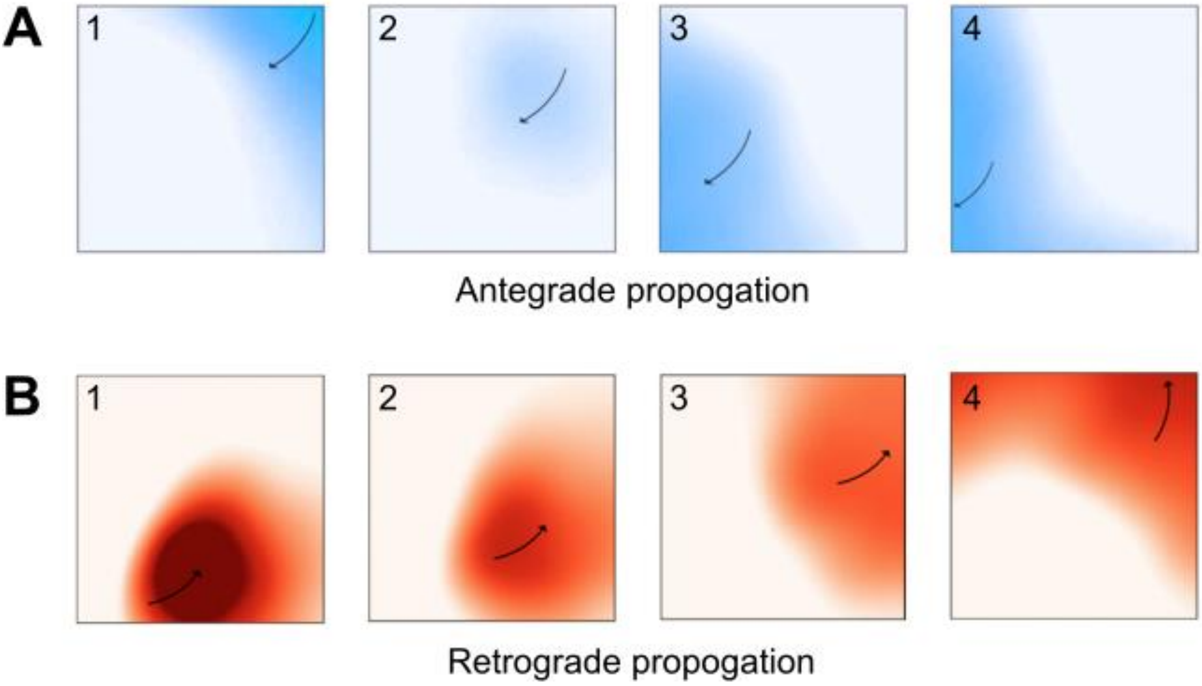
